# Association of patients’ past misdiagnosis experiences with trust in their current physician: the TRUMP^2^-Net study

**DOI:** 10.1101/2021.01.25.21250300

**Authors:** Ryo Suzuki, Nobuyuki Yajima, Kosuke Sakurai, Nao Oguro, Takafumi Wakita, David H. Thom, Noriaki Kurita

## Abstract

**Background:** Previous qualitative research has described that past misdiagnosis experiences may reduce patients’ own and their families’ trust in healthcare.

**Objective:** To quantify the associations between patients’ or family members’ misdiagnosis experiences and the former’s trust in their current physicians.

**Design:** A cross-sectional online survey.

**Participants:** Adult Japanese people with non-communicable diseases (cancer, diabetes, depression, heart disease, and connective tissue disease), recruited using a web-based panel survey.

**Main Measures:** The misdiagnosis experiences of patients and their family members were measured as exposures. The former’s trust in their current physicians was measured using the Japanese version of the 11-item Trust in Physicians Scale modified by Thom, which was translated and validated by us for this study.

**Key Results:** A total of 661 patients with a mean age of 62.7 years were analyzed. Overall, 23.2% had a history of misdiagnosis and 20.4% had a family member who had been misdiagnosed. The internal consistency (Cronbach’s α) was 0.91. The factor analysis suggested unidimensionality with all 11 loadings being higher than 0.40. In a multivariable-adjusted general linear model, patients’ and family members’ misdiagnosis experiences were associated with lower confidence in their current physicians (mean difference −4.30, 95%CI −8.12 to −0.49 and −3.20, 95%CI −6.34 to −0.05, respectively). An additive effect was suggested for the associations of patients’ and family’s experience of misdiagnosis on trust (P for interaction = 0.494).

**Conclusions:** The individuals’ and family members’ misdiagnosis experiences were associated with reduced trust in their current physicians. Interventions specifically targeting misdiagnosed patients are needed to restore patients’ confidence in their current physicians.

## Introduction

Patient trust in physicians is central to patient-physician relationship.^1^ It refers to patients’ belief in the physician’s credibility or their confidence in the latter’s capacity to influence health outcomes, called competence.^2^ Its importance is highlighted in the shared decision-making context, as interpersonal trust between patients and clinicians fosters shared decision-making regarding treatment plans.^3^ Loss of trust in physician negatively affects health behaviors.^4, 5^ Additionally, misdiagnosis is a serious cause, as the patient’s negative experience diminishes perceptions of physician’s competence, which is a theoretical component of trusting them. Approximately 5% of the general population in the United States are misdiagnosed, which accounts for about a third of preventable deaths^7^ and leads to medical litigation.^8, 9^ Despite the serious consequences, limited studies have examined the impact of previous misdiagnosis experiences on trust in future physicians.

Previously, the effects of misdiagnosis experiences on the trust of patients and their families were described as components of medical errors. Qualitative studies report that medical errors, including misdiagnosis, cause patients and their families to lose confidence in healthcare and avoid medical care,^10^ which persist for over five years.^11^ Although misdiagnosis remains a vivid negative memory for patients^11^ and may shape their attitudes toward other physicians,^12^ no quantitative studies have assessed whether patients’ or their family members’ past misdiagnosis experiences affect their trust in their current physicians. Studies quantifying such impacts could help to develop interventions to restore trust in current physicians and maintain a favorable therapeutic relationship.

Therefore, this study investigated the associations of Japanese patients’ and their family members’ misdiagnosis experiences with trust in their current physicians by analyzing data from an online survey: the Trust Measurement for Physicians and Patients–the Net survey (the TRUMP^2^-Net).

## Methods

### Setting and participant selection

The TRUMP^2^-Net study, a cross-sectional online survey, was approved by the Institutional Review Board at Kansai University. We conducted a panel survey supported by a web-based company (Cross Marketing, Shinjuku-ku, Tokyo) to recruit Japanese patients with non-communicable diseases, aged 20 years or older. Participants were offered an incentive and points. In principle, participants were prohibited from answering more than once, and researchers could only use the initial entry if they answered more than once. Only those who accepted an informed consent statement could answer the questionnaire. As with the Amazon Mechanical Turk in the United States, a response rate could not be calculated for this survey.

### Designing screener items

As this was a web-based survey, the presence of careless participants cannot be ignored.^13, 14^ To prevent random variability and reliability loss in the answers, multiple “screener” items were designed to exclude them from our analysis.^14^

Screening for non-communicable diseases

First, participants were asked to select one of the following diseases for which they had received medical treatment twice or more within the past six months: heart disease (arrhythmia), heart disease (angina pectoris, myocardial infarction), heart disease (heart failure), diabetes, rheumatic diseases (rheumatoid arthritis), rheumatic diseases (systemic lupus erythematosus), cancer (limited only to those currently being treated), and depression. Multiple choices were allowed. Then, they were instructed to choose the illness that was most troublesome from those chosen previously. Participants who selected a disease different from the previously-chosen item were excluded.

### Validation by self-reported drug name

Participants were instructed to provide the name of a medication prescribed for the most troubling disease in a free-text format. We searched for label information online to assess whether the relevant disease was listed for the indication, in which case, the responses were considered valid. Those who chose cancer and answered “none” for their prescribed drugs were also included, as not all cancer treatments require drug prescriptions. Two researchers conducted these assessments independently; if the evaluations varied, decisions were reached through discussions.

### Response time

Participants were screened using a cutoff response time because those who responded too quickly could be careless.^13, 14^ We measured response time for five researchers and two research assistants prior to the panel survey and found that at least five minutes (300 seconds) were required to complete the survey. Therefore, those taking under 300 seconds were excluded.

### Modified version of the Trust in Physician Scale

For this study, the 11-item Trust in Physician Scale modified by Thom,^4^ was translated into Japanese after obtaining the original developer’s permission. Two physicians (N.Y. and N.O.), a physician researcher (N.K.), and a quantitative psychologist (T.W.) with experience in scale development translated the scale into Japanese. Then, it was back-translated into English by two bilingual individuals (one American and one Canadian). The items were compared with the original items, and the translated and back-translated versions were amended. Finally, they were sent to the original author, and minor improvements were made. The final version was approved by the original author (Supplementary Table 1).

**Table 1.** Participant characteristics (N = 661)

|  | n (%) |
| --- | --- |
| **Age, in years<sup>a</sup>** | 62.7 (10.1) |
| **Female, n (%)** | 175 (26.5%) |
| **Education, n (%)** |  |
| *Junior high school* | 19 (2.9%) |
| *High school* | 209 (31.6%) |
| *Junior college* | 65 (9.8%) |
| *University* | 325 (49.2%) |
| *Graduate school* | 30 (4.5%) |
| *Not answered* | 13 (2.0%) |
| **Total household income, n (%)** |  |
| *< 1,000,000 yen* | 40 (6.1%) |
| *1,000,000 – < 3,000,000 yen* | 157 (23.8%) |
| *3,000,000 – < 5,000,000 yen* | 203 (30.7%) |
| *5,000,000 – < 10,000,000 yen* | 205 (31.0%) |
| *≥ 10,000,000 yen* | 56 (8.5%) |
| **Region, n (%)** |  |
| *Hokkaido* | 35 (5.3%) |
| *Tohoku* | 34 (5.1%) |
| *Chubu* | 98 (14.8%) |
| *Kanto* | 273 (41.3%) |
| *Kansai* | 132 (20.0%) |
| *Chugoku* | 29 (4.4%) |
| *Shikoku* | 19 (2.9%) |
| *Kyushu-Okinawa* | 41 (6.2%) |
| **Reported disease, n (%)** |  |
| *Cardiac disease, arrhythmia* | 37 (5.6%) |
| *Cardiac disease, angina pectoris or myocardial infarction* | 119 (18.0%) |
| *Cardiac disease, heart failure* | 15 (2.3%) |
| *Diabetes* | 191 (28.9%) |
| *Connective tissue disease* | 17 (2.6%) |
| *Cancer* | 255 (38.6%) |
| *Depression* | 127 (19.2%) |
| **The most troublesome disease, n (%)** |  |
| *Cardiac disease, arrhythmia* | 17 (2.6%) |
| *Cardiac disease, angina pectoris or myocardial infarction* | 89 (13.5%) |
| *Cardiac disease, heart failure* | 8 (1.2%) |
| *Diabetes* | 175 (26.5%) |
| *Connective tissue disease* | 13 (2.0%) |
| *Cancer* | 242 (36.6%) |
| *Depression* | 117 (17.7%) |
| **Duration of relationship with physician, n (%)** |  |
| *< 1 year* | 60 (9.1%) |
| *1 – <3 year* | 212 (32.1%) |
| *≥ 3 year* | 389 (58.9%) |

Continuous variables summarized as means and standard deviations (in parentheses).
Categorical variables summarized as frequencies and proportions (in square brackets).

Before answering the questionnaire, participants read the following statements: “Please answer about your doctor who provides care for your [the most troublesome disease selected by the participant was automatically displayed here]. How much do you agree or disagree with the following statements?” Participants were instructed to rate each item on a Likert scale ranging from 1 (*totally disagree*) to 5 (*totally agree*). The sum of all item scores was converted to that of 0-100.^4^

### Experience with misdiagnosis

To assess participants’ own experience of misdiagnosis, they were first given the following instruction: “Looking back at all the medical care you have received, please choose 1 (*have had*) or 2 (*have not*) if you have or have not experienced it, respectively.” Then, the following question was asked: “Have you been misdiagnosed about your illness?”

To assess a family member’s misdiagnosis experience, participants were provided the same aforementioned instructions, following which, the question “Has your family member ever been misdiagnosed by a physician?” was asked.

### Other patient survey variables

A detailed description of items selection and their hypothesized correlations with patient trust are provided in Supplementary Item 1. Patient satisfaction, scored on a 5-point Likert scale ranging from *strongly disagree* to *strongly agree*, was assessed using the item, “Overall, you are extremely satisfied with the doctor?”^15^ Patients’ willingness to follow their physicians, scored on a 5-point Likert scale ranging from *strongly disagree* to *strongly agree*, was examined using the item, “If my doctor moves to another medical institution, I would like to visit that institution to see them.”^16^ Further, physicians’ supportive attitudes during visits, scored on a 6-point Likert ranging scale from *completely disagree* to *completely agree*, were assessed using the item, “My doctor helped me understand all the information.”^17^ This item is considered as an attitude that reflects patients’ trust in their physician to prioritize what the patient cares about and provide required medical support.^2^ The item “I can overcome most illnesses without a physician’s help” examined patients’ attitude toward medical care; it was scored on a 5-point Likert scale ranging from *strongly disagree* to *strongly agree*.^18^ This is a modified item from a scale that evaluates medical skepticism.^19^ Patients’ general level of interpersonal trust was assessed using the 6-item General Trust Scale,^20^ rated on a 5-point Likert-type scale ranging from *strongly disagree* to *strongly agree*. The score was computed by summing the items.

Demographic characteristics including age, sex, education level, total household income, and zip code were collected as covariates. We categorized residents’ prefectures based on the first three digits of the zip code. The durations of the patient-physician relationships were categorized as less than 1 year, 1–3 years, and more than 3 years.

### Statistical analysis

All statistical analyses were performed using the Stata/SE version 15 (Stata Corp., College Station, TX, USA). Participant characteristics were summarized as mean and standard deviation for continuous variables, and frequencies and proportions for categorical variables.

A factor analysis using the MINRES examined the factorial structure of the Trust in Physician Scale items. The number of latent factors was assessed by the eigenvalue attenuation.^21^ The absolute factor loadings were calculated. We performed a confirmatory factor analysis to evaluate the fit of the one-factor model to the data, because the original scale had one factor.^4^ Reliability was assessed using the Cronbach’s α and the McDonald’s ω coefficients.^22^ Construct validity was examined by using the correlation between the Trust in Physician Scale and the following factors: patient satisfaction, patient’s choice of physician, physician’s supportive attitude during the visit, and duration of patients’ relationship with their physician.^4^ Additionally, it was assessed by testing the correlation between the Trust in Physician and the General Trust scales and patients’ attitude toward medical care. Spearman’s correlation coefficient was calculated to test the correlation.

To estimate the association between misdiagnosis experience and trust in physicians, we fitted a series of general linear models with cluster-robust variance that accounted for the clustering effect by prefectures. In unadjusted analyses, patients’ and their family’s misdiagnosis experiences were fit to a separate model. In the multivariable-adjusted analysis, patients’ and their families’ experience of misdiagnosis, as well as covariates (age, gender, level of education, total household income, comorbidities, and the duration of the relationship with their physicians), were fitted to a single model. These covariates were chosen as they could be associated with both trust in physicians and the misdiagnosis experience. To examine any interaction between patients’ and their families’ misdiagnosis experience, their product term was entered into the multivariable-adjusted model. Interaction was assessed using the Wald test.

## Results

Overall, 964 individuals participated in the study (Figure 1). After excluding 293 and 10 individuals due to three screen items and missing covariates, respectively, 661 were included in the primary analysis.

**Figure 1.**
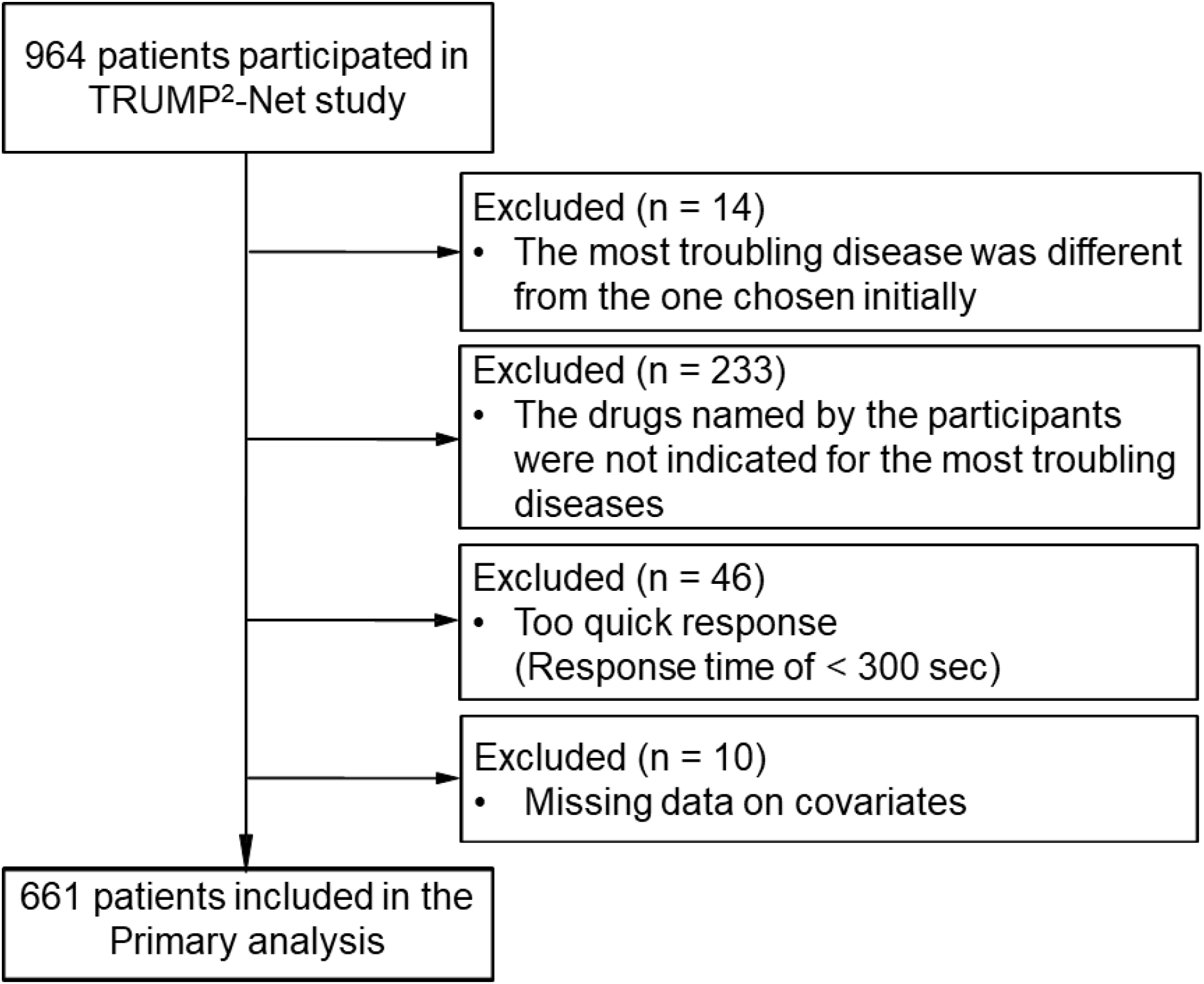
Flow of the study. Abbreviations: TRUMP^2^-Net, Trust Measurement for Physicians and Patients - the Net survey

### Participant Characteristics

Participant characteristics are presented in Table 1. The mean age was 62.7 years, and 175 (26.5%) participants were female. The region of residence extended to 46 prefectures, with Kanto being the most common one (41.3%), followed by Kansai (20.0%). Cancer was the most common troublesome disease (36.6%), followed by diabetes (26.5%), depression (17.7%), and heart disease (17.3%).

Descriptive statistics and psychometric testing of the Trust in Physician Scale

The eigenvalue attenuation indicated that the items had a strong unidimensionality (Supplementary Figure 1). The absolute values of factor loadings for each item suggested that each item could be included in a single factor (Supplementary Table 2). The confirmatory factor analysis resulted in a root mean square error of 0.086, a comparative fit of index of 0.95, and a standardized root mean square residual of 0.049, indicating a good fit of the one-factor model to the data. Cronbach’s alpha and the McDonald’s ω coefficients were 0.91 and 0.93, respectively.

**Table 2.** Correlation between the Trust in Physician Scale and selected variables.

|  | **Correlation**<br>**coefficients<sup>a</sup>** | ***P*-value** |
| --- | --- | --- |
| Patient's satisfaction with the physician | 0.732 | < 0.001 |
| Patients' willingness to follow their physician | 0.347 | < 0.001 |
| Physician's supportive attitudes during visits | 0.731 | < 0.001 |
| Duration of the relationship with the physician | 0.078 | 0.044 |
| Patient's attitude toward medical care | -0.205 | < 0.001 |
| Patient's general level of interpersonal trust | 0.300 | < 0.001 |

<sup>a</sup>All variables were tested using Spearman's correlation coefficient.

The mean score was 70.5 (standard deviation: 15.1). The scores were distributed from 11.4 to 100, with only 2.6% of participants at the ceiling score of 100. The Trust in Physician Scale strongly correlated with satisfaction with the physician (ρ = 0.732) and their supportive attitudes during the visit (ρ = 0.731), confirming its construct validity (Table 2). Furthermore, the scale weakly correlated with the willingness to follow their physicians (ρ = 0.347) and very weakly with the duration of the relationship with the physician (ρ = 0.078). It showed a weak positive correlation with general interpersonal trust (ρ = 0.300), suggesting that it measured a different concept. Furthermore, it was weakly and negatively correlated with skeptical attitudes toward medical care (ρ = −0.205).

### Frequency of misdiagnosis experiences and its relationship with trust in physician

Overall, 153 participants (23.2%) had a history of misdiagnosis, and 135 (20.4%) had a family member who had been misdiagnosed. Of all the participants, 71 (10.7%) had been misdiagnosed and had a family member who had experienced the same.

The association between misdiagnosis experience and trust in physicians is shown in Table 3. There is insufficient evidence to suggest the joint effect of the individual’s and the family’s misdiagnosis experience on trust (P for interaction = 0.494); therefore, analyses were performed without considering the interaction term between the two. Both the former (mean difference −4.30, 95%CI −8.12 to −0.49) and the latter were associated with lower trust (mean difference: −3.20, 95%CI −6.34 to −0.05). The individuals’ and family’s misdiagnosis experiences were additively associated with lower trust compared to neither (Figure 2: Mean difference: −7.50, 95%CI −10.5 to −4.53).

**Table 3.** Associations between misdiagnosis experience and the Trust in Physicians.

| Trust in Physicians Scale, points | Mean difference | (95% CI) | P-value |
| --- | --- | --- | --- |
| **<u>Unadjusted</u>** |  |  |  |
| Patient's experience with a misdiagnosis | **-5.80** | **(-9.09 to -2.51)** | **0.001** |
| Patient's family experience with a misdiagnosis | **-5.19** | **(-8.23 to -2.14)** | **0.001** |
| **<u>Multivariable-adjusted<sup>b</sup></u>** |  |  |  |
| Patient's experience with a misdiagnosis | **-4.30** | **(-8.12 to -0.49)** | **0.028** |
| Patient's family experience with a misdiagnosis | **-3.20** | **(-6.34 to -0.05)** | **0.047** |
| Age, per 10 years | **1.17** | **(0.06 to 2.29)** | **0.040** |
| Sex, female | 1.17 | (-1.49 to 3.83) | 0.379 |
| Duration with patients' physician |  |  |  |
| < 1 year | **-4.28** | **(-8.3 to -0.28)** | **0.036** |
| 1 – < 3 years | **-3.15** | **(-5.5 to -0.76)** | **0.011** |
| ≥ 3 years | Ref |  |  |
| Education |  |  |  |
| Junior high school | Ref |  |  |
| High school | -1.34 | (-6.6 to 3.88) | 0.607 |
| Junior college | 1.48 | (-5.6 to 8.58) | 0.677 |
| University | -0.43 | (-5.7 to 4.84) | 0.869 |
| Graduate school | -3.67 | (-12.0 to 4.61) | 0.377 |
| Not answered | -1.16 | (-9.2 to 6.85) | 0.772 |
| Total household income |  |  |  |
| < 1,000,000 yen | **-9.22** | **(-15.1 to -3.38)** | **0.003** |
| 1,000,000 - < 3,000,000 yen | -3.29 | (-8.22 to 1.64) | 0.186 |
| 3,000,000 - < 5,000,000 yen | -4.79 | (-9.89 to 0.31) | 0.065 |
| 5,000,000 - < 10,000,000 yen | -4.42 | (-9.69 to 0.86) | 0.099 |

| $\geq 10,000,000$ yen | Ref |
| --- | --- |
| Analysis of 661 patients from 46 prefectures. |  |
| <sup>a</sup>General linear models with consideration of prefectural level correlation using cluster-variance. |  |
| <sup>b</sup>General linear model adjusted for age, sex, duration of the relationship with the physician, comorbidities, education, and total household income with consideration of prefectural level correlation using cluster variance. The variable in each unadjusted model was included in the multivariable model. |  |

**Figure 2.**
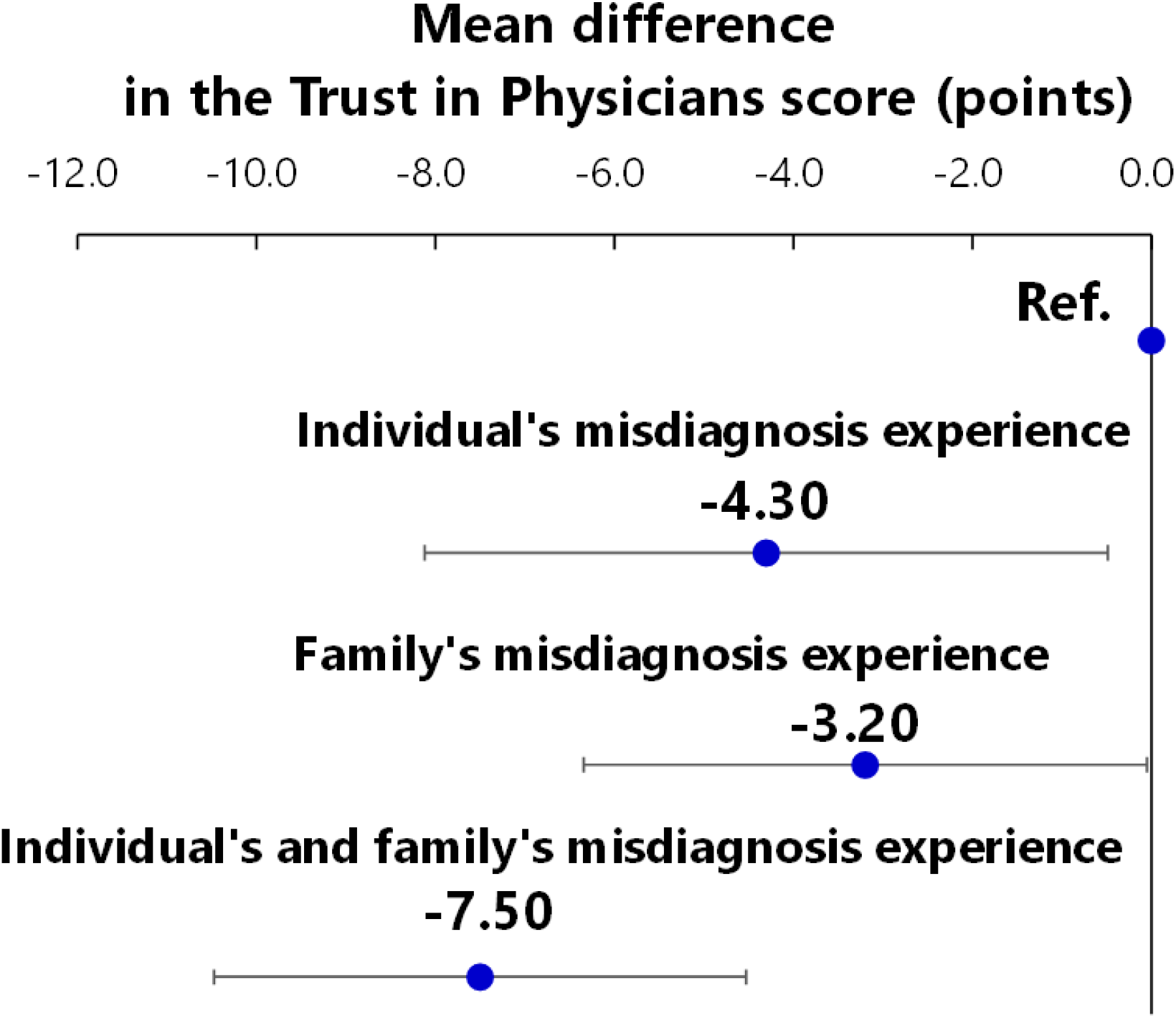
Associations of individual and family misdiagnosis experiences with the Trust in Physicians score. Mean differences were estimated using a general linear model adjusted for age, sex, duration of patient-physician relationship, comorbidities, education, and total household income with consideration of prefectural-level correlation using cluster variance.

Older participants had higher trust scores than younger ones (mean difference per 10-year difference: 1.17, 95%CI: 0.06–2.29). A total household income of less than 1 million yen was associated with lower trust compared to household income over 10 million yen (mean difference: −9.22, 95%CI: −15.1 to −3.38). Additionally, a shorter relationship with the physician was associated with lower trust (mean difference for less than 1 year and for 1–3 years versus more than 3 years: −4.28, 95%CI: −8.27 to −0.28 and −3.15, 95%CI: −5.54 to −0.76, respectively).

## Discussion

In this study, both patients’ and family members’ past misdiagnosis experiences were associated with lower levels of patients’ trust in their current physicians. Additionally, patients’ and family members’ past misdiagnosis experiences had almost a two-fold additive effect on the former’s trust in their current physicians. These findings encourage physicians to pay attention to overlooked sources of distrust and implement new strategies in outpatient settings to restore trust of patients with non-communicable diseases.

Our novel findings also support previous qualitative findings about patients’ and their family members’ negative emotions and behavioral responses to healthcare post misdiagnosis, and confirm universal behaviors of patients’ trust in physicians, as revealed in previous research. First, we were found that loss of trust extends to current physicians after misdiagnoses, while prior qualitative studies found it in physicians who had made medical errors.^10, 11^ Considering the vivid, negative emotions and persisting memories from past medical errors including misdiagnosis,^11^ loss of trust in current physicians is reasonable, and this finding supports the theory that past experiences shape patients’ attitudes toward new physicians.^12^ Second, unlike previous studies that treated medical errors and misdiagnosis as components,^10, 11, 23^ we focused on misdiagnosis among medical errors. Since diagnostic accuracy can be considered part of a physician’s competence, a misdiagnosis may make patients lose confidence in their physicians and skeptical of other physicians.^24^ Third, this study is the first to investigate additive associations of patients’ and their families’ misdiagnosis experiences with the former’s trust in their current physicians. Our findings suggest the additive effect of memories from negative and painful experiences. Fourth, our findings suggesting the association of higher levels of trust in physicians with being older and higher income levels are consistent with those of previous studies conducted in the United States,^4, 25^ and indicate that interpersonal trust in physicians is a basic cognitive activity that is similar across countries.

Our findings have implications for physicians and researchers. First, physicians can inquire about patients’ and family members’ misdiagnosis experiences as part of routine medical history taking. Second, patients with a family history of misdiagnosis, in addition to their own, may have lower levels of trust; they should be assessed for the need to reconstruct trust. However, thus far, there is insufficient evidence for specific effective interventions aimed at rebuilding trust in physicians.^26^ Developing appropriate interventions, such as training programs for physicians to learn communication skills and patient education, are of clinical importance. Such training could include communication skills, such as listening to the timing and nature of the misdiagnosis, the consequent emotions, psychological distress; sympathizing with the patient’s attitudes toward and concerns about the current medical care they are receiving; and identifying with the patient the benefits of the current medical care.

This study has several strengths. First, our findings may apply to various settings as it included patients with chronic conditions. Second, our results were likely unaffected by geographical differences because the possibility of prefectural-level difference in trust was addressed by statistical modeling using a cluster-robust estimation, although previous studies suggest regional differences in residents’ trust in others.^25, 27^ As our survey targeted patients in Japan who could answer our Japanese-language questionnaire, all participants were assumed to be Japanese. Therefore, observed associations were likely unaffected by racial differences.

Several limitations of this study warrant mention. First, the sample may not be representative of patients with the same non-communicable diseases. In fact, men were about three times more represented than women. However, this likely does not affect the findings, as gender was adjusted for in the analyses.Second, we were unable to determine whether the misdiagnosis experience was related to a non-communicable disease currently being treated or to a past one.

In conclusion, individuals’ and family members’ misdiagnosis experiences were associated with reduced trust in current physicians. Furthermore, these misdiagnosis experiences had an additive effect. Future studies should develop interventions to restore lost confidence from past misdiagnosis experiences.

## Data Availability

The datasets used and/or analyzed during the current study are available from the corresponding author on reasonable request.

## Acknowledgments

The abbreviated name of the study, “the TRUMP^2^-Net (the Trust Measurement for Physicians and Patients - the Net survey)” was not derived from a specific individual, however, it was determined to suggest a trusted physician, through consideration of the fact that the word “trump” has an ancient meaning of “a dependable and exemplary person.” This study was supported by the JSPS KAKENHI (Grant Number: JP 19KT0021). The funders had no role in the study design, analysis, or interpretation of the data; writing of the manuscript; or the decision to submit it for publication.

