## Supplemental file for "Association of patients’ past misdiagnosis experiences with trust in their current physician: the TRUMP^2^-Net study"

### Supplementary Table 1. Japanese version of the Trust in Physicians Scale

| Instruction | 以下の文章について、あなたはどの程度そう思いますか。最もよく表している回答の番号に○をつけてください。*  (Original: “Please circle the number for the response that best describes how much you agree or disagree with each of the following statements.”) |
| --- | --- |
| Question 1 | 主治医は私を一人の人間として気にかけてくれているようには思わない。  (Original: “I doubt that my doctor really cares about me as a person.”) |
| Question 2 | 主治医はふだんから私の要望に配慮して、第1に考えてくれる。  (Original: “My doctor is usually considerate of my needs and puts them first.”) |
| Question 3 | 私は主治医をとても信頼しているので，アドバイスには常にしたがおうと思う。  (Original: “I trust my doctor so much I always try to follow his/her advice.”) |
| Question 4 | 主治医が私に伝えることは、常に真実のはずだ。  (Original: “If my doctor tells me something is so, then it must be true.”) |
| Question 5 | 私はときどき主治医の意見が信頼できず，別の医師の意見を聞きたいと思う。  (Original: “I sometimes distrust my doctor's opinion and would like a second one.”) |
| Question 6 | 私は，自分の治療に関して主治医の判断を信じている。  (Original: “I trust my doctor's judgments about my medical care.”) |
| Question 7 | 私は, 主治医が必要な全ての治療を行なってくれていないと感じる。  (Original: “I feel my doctor does not do everything he/she should for my medical care.”) |
| Question 7 | 私は, 主治医が必要な全ての治療を行なってくれていないと感じる。  (Original: “I feel my doctor does not do everything he/she should for my medical care.”) |
| Question 8 | 主治医が私の病気を診るときは，治療に必要なことを最優先してくれていると信じている。  (Original: “I trust my doctor to put my medical needs above all other considerations when treating my medical problems.”) |
| Question 9 | 主治医は私のような病気に対処（診断，治療，適切な紹介をする）する能力が十分にあると思う。  (Original: “My doctor is well qualified to manage (diagnose and treat or make an appropriate referral) medical problems like mine.”) |
| Question 10 | 私の治療に間違いがあれば, 主治医は私に伝えてくれると信じている。  (Original: “I trust my doctor to tell me if a mistake was made about my treatment.”) |
| Response options for Question | 全くそう思わない/そう思わない/どちらともいえない/そう思う/とてもそう思う  (Original: totally disagree/disagree/neutral/agree/totally agree) |

The original English version^1^ is also provided for each item and response.

Before using this instrument, please register at https://noriaki-kurita.jp/resources/trust-in-physician-jpn/.

In addition, please cite this article as follows:

Suzuki R, Yajima N, Sakurai K, Oguro N, Wakita T, Thom DH, Kurita N. Association of patients’ past misdiagnosis experiences with trust in their current physician: the TRUMP^2^-Net study. *J Gen Intern Med*. 2021.

*The instructional statements generated through the formal process of translation are presented. Modified instructional statements were used in the online survey of this study as follows: “Please answer about your doctor who cares for your [the most troublesome disease selected by the participants was automatically displayed in this place]. How much do you agree or disagree with each of the following statements.”

### Supplementary Item 1. A detailed description of items selection and their hypothesized correlations with the Trust in Physician Scale

Patient satisfaction, scored on a 5-point Likert scale ranging from 1 (*strongly disagree*) to 5 (*strongly agree*), was assessed using the item, “Overall, you are extremely satisfied with the doctor.”^1^ Previous studies have reported strong correlations between patient satisfaction and trust in the physician.^1-3^ Patients’ willingness to follow their physicians, scored on a 5-point Likert scale ranging from 1 (*strongly disagree*) to 5 (*strongly agree*), was examined using the item, “If my doctor moves to another medical institution, I would like to visit that institution to see them.”^4^ Given that confidence and reassurance in one’s physician and their intentions indicate trust in their physician,^5^ a higher interpersonal trust score indicated a stronger desire to continue being treated by that physician. Furthermore, physicians’ supportive attitudes during visits, scored on a 6-point Likert ranging scale from 0 (*completely disagree*) to 5 (*completely agree*), were assessed using the item, “My doctor helped me understand all the information.”^6^ This item is part of the Japanese version of the 9-Item Shared Decision-Making Questionnaire,^6^ and can be considered as an attitude that reflects patients' trust in their physician to prioritize what the patient cares about and will provide required medical support.^3^ The item “I can overcome most illnesses without a physician’s help” assessed patients’ attitude toward medical care; it was scored on a 5-point Likert scale ranging from 1 (*strongly disagree*) to 5 (*strongly agree*).^7^ This item is a modified version of an item from a scale that evaluates medical skepticism.^8^ We modified the word “medically trained professional” to “physician” because only the latter is allowed to provide medical care in Japan. It is assumed that the more the participants prefer self-care over physician control, the less they trust their physicians.^3^ Patients’ general level of interpersonal trust was assessed using the 6-item General Trust Scale^9^ rated on a 5-point Likert-type scale ranging from 1 (*strongly disagree*) to 5 (*strongly agree*). The total score was computed by summing the item scores. Although the Trust in Physician Scale is expected to be related it, this association is weak because it measures trust in a specific patient-physician relationship rather than the general interpersonal trust.^2^

### Supplementary Figure 1. Scree plot for the eigenvalues using responses to the Trust in Physician Scale

The eigenvalue attenuation was the largest between the first and second factors (6.19, 1.09, and 0.71 for the first, second, and third factors, respectively), indicating that the items had a strong unidimensionality.

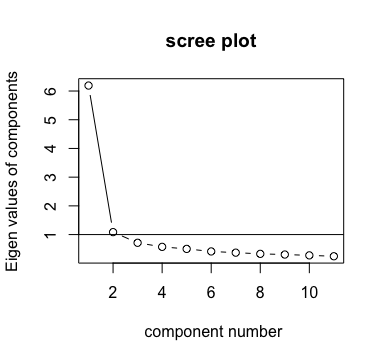

### Supplementary Table 2. Descriptive statistics for the Trust in Physician Scale’s items

| Item No. | Question | Mean | SD | Factor loading |
| --- | --- | --- | --- | --- |
| 1 | 主治医は私を一人の人間として気にかけてくれているようには思わない。 | 2.35 | 0.90 | -0.63 |
| 2 | 主治医はふだんから私の要望に配慮して、第1に考えてくれる。 | 3.62 | 0.78 | 0.76 |
| 3 | 私は主治医をとても信頼しているので，アドバイスには常にしたがおうと思う。 | 3.75 | 0.76 | 0.81 |
| 4 | 主治医が私に伝えることは、常に真実のはずだ。 | 3.72 | 0.78 | 0.75 |
| 5 | 私はときどき主治医の意見が信頼できず，別の医師の意見を聞きたいと思う。 | 2.47 | 0.96 | -0.69 |
| 6 | 私は，自分の治療に関して主治医の判断を信じている。 | 3.87 | 0.75 | 0.86 |
| 7 | 私は, 主治医が必要な全ての治療を行なってくれていないと感じる。 | 2.45 | 0.99 | -0.48 |
| 8 | 主治医が私の病気を診るときは，治療に必要なことを最優先してくれていると信じている。 | 3.77 | 0.75 | 0.83 |
| 9 | 主治医は私のような病気に対処（診断，治療，適切な紹介をする）する能力が十分にあると思う。 | 3.85 | 0.78 | 0.81 |
| 10 | 私の治療に間違いがあれば, 主治医は私に伝えてくれると信じている。 | 3.72 | 0.76 | 0.79 |
| 11 | 私は，主治医と話した内容を二人だけの秘密にしていないと, ときどき心配に思う。 | 2.19 | 0.84 | -0.42 |

The absolute values of the factor loadings for each item ranged from 0.42 to 0.86. Thus, each item could be included in a single factor.
